# A Simple Method for Estimating the Number of Unconfirmed COVID-19 Cases in a Local Area that Includes a Confidence Interval: A Case Study of Whatcom County, Washington

**DOI:** 10.1101/2020.04.30.20086181

**Authors:** David A. Swanson, Ronald E. Cossman

## Abstract

Along with many other data problems affecting the unfolding of the COVID-10 pandemic in the United States, virtually nothing is known about the number of positive, unconfirmed cases, especially in local areas. We show that it is possible to estimate the number of positive, unconfirmed COVID-19 cases using a simple, long-established method employed by demographers to estimate a population in the absence of a census count. We go on to show how a confidence interval can be constructed around an estimate of positive, unconfirmed COVID-19 cases constructed from this method, using Whatcom County, Washington as a case study.

## Introduction

Along with many other data problems affecting the unfolding of the COVID-10 pandemic in the United States, virtually nothing is known about the number of positive, unconfirmed cases, especially in local areas, In this paper, we show that it is possible to estimate the number of positive, unconfirmed COVID-19 cases using a simple, long-established method employed by demographers to estimate a population in the absence of a census count, known as the censal-ratio method (Swanson and Tayman, 2012: 187-194). We go on to show how a confidence interval can be constructed around such an estimate, using Whatcom County, Washington as a case study.

As a roadmap to the development of estimates for Whatcom County, Washington, we start by combining information from two sources. The first is an estimate of the “case fatality rate” from a study in Germany of the municipality of Gangelt (Bailey, 2020). The second source is information for the U.S. as a whole provided by the Centers for Disease Control (https://data.cdc.gov/NCHS/Provisional-Death-Counts-for-Coronavirus-Disease-C/hc4f-j6nb/data), including the number of confirmed COVID-19 cases and the number of deaths due to COVID-19 among those same confirmed cases, assembled around April 9^th^.

From these two sources, we then estimate the total number of positive, unconfirmed COVID-19 for the U.S. as a whole by dividing the U.S. COVID-19 deaths by the by the Gangelt Case Fatality Rate. We divide this estimate by the number of confirmed COVID-19 cases for the U.S. as a whole to yield a “multiplier,” the number of positive, unconfirmed cases relative to the number of confirmed cases for the U.S. as a whole. We then apply standard measure used in inferential statistics to obtain an estimate of the 95% confidence boundaries for the U.S. multiplier. We conclude with the application of these steps to Whatcom County, Washington to obtain an estimate of its number of positive, unconfirmed cases along with 95% lower and upper confidence limits.

## Methods and Data

The censal-ratio method can be implemented in several different ways (Swanson and Tayman, 2012: 197-194). The most basic approach is to use relationships between symptomatic indicators and population counts in census years to estimate populations in non-census years and applying these relationships to symptomatic indictors available in the years for which estimates are desired. The general form of this approach is as follows.

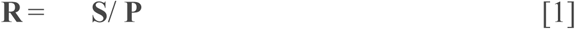

where

R = censal-ratio
P = population
S = symptomatic indicator (e.g., recorded deaths, births, school enrollment)

Once a censal-ratio is constructed, a population estimate for area i is developed by dividing the value of the symptomatic indicator (S_i_) by the ratio (R) to yield an estimate of *P*_i_:

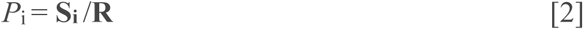

One advantage of using Equation [1] is that the resulting ratio of interest is easy to interpret. If one uses deaths as the symptomatic indicator, then the ratio is the crude death rate. Similarly, if one uses births, the resulting ratio is the crude birth rate.

In adapting the censal-ratio method to estimate the number of positive, unconfirmed COVID-19 cases, we revise equations [1] and [2] as follows

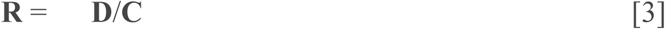

where

**R** = Ratio of deaths due to COVID-19 to the Total number of COVID-19 cases found in Gangelt.
**D** = deaths due to COVID-19 in Gangelt
**C** = Total number of COVID-19 cases found in the Gangelt “sample”

Note that in this case, **R** is equal to the Case Fatality Rate for Gangelt.

Once **R** is found, it can be used to estimate the number of positive, unconfirmed cases, *U_i_*, for a given area, i, by dividing it into the reported deaths due to COVID-19 in area i, **D_i_**

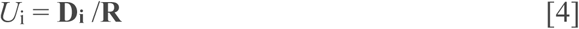

As we illustrate below, once *U*_i_ is estimated, we also can find the ratio of unconfirmed cases (*U*_i_) to confirmed cases (**C**_i_), viz., ratio = *U*_i_/ **C**_i_

This ratio is useful, first, because we can use it to estimate the number of unconfirmed cases in a given area of interest, w (*U*_w_), by multiplying it by the confirmed cases in area w, **C_w_**

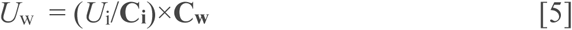

The ratio useful for a second reason, which is that it makes the process of developing a confidence interval around an estimate of *U*_w_, a process we will return to shortly.

Here is a concrete example of these steps using the data described earlier. By dividing the case fatality rate found in the German study (**R** = 0.004), into the number of COVID-19 deaths in the U.S. (**D_i_** = 18,559), we obtained an estimate of *U*_i_ = 4,639,790 unconfirmed positive cases in the U.S.

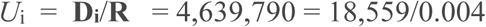

### Results: Estimated Positive, Unconfirmed Cases

In order to apply this estimate to any given area, we then divide this number by the number of confirmed COVID-19 cases in the U.S. provided by CDC (492,416) to find the ratio of unconfirmed to confirmed cases, which yields *U*_i_/ **C**_i_ = 9.4, where 9.4 = 4,639,790/492,416.

By multiplying 9.4, by the number of confirmed cases in a given area for which they are reported, one obtains an estimate of its positive, unconfirmed cases. Applying this to Whatcom County, Washington which as of April 25^th^ reported 284 confirmed cases (Johns Hopkins University, 2020), we estimate that there were 2,670 unconfirmed, positive cases on that date.

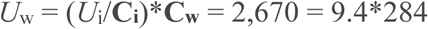

One issue with this approach is that some of the confirmed cases are no longer active, due to death or recovery. Applying this method to the number of active cases (257) reported for Whatcom County, Washington as of April 25^th^ (Johns Hopkins University Coronavirus Resource Site, 2020) yields an estimated 2,416 positive, unconfirmed cases, where *U*_w_ = 2,416 = 9.4^*^ 257.

### Developing a Confidence Interval

Swanson and Tayman (2012: 189-191) describe how inferential measures such as the coefficient of variation can be obtained from censal-ratio methods. Extending this to our example of the estimated number of unconfirmed cases in the United States (4,639,790), we start with the German study (Bailey, 2020), which took place in Gangelt, a municipality with a 2018 population of 12,446 (https://en.wikipedia.org/wiki/Gangelt). In the study, 80 percent of this population was tested (Bailey, 2020), which yields a study “sample” of approximately 9,957. While this was not a random sample in the sense of survey research, we can take the view that this is random sample from a “super population” of infinite possible outcomes (Hartley and Sielken 1975; Sampath 2005). Recall that the case fatality rate in the German study is 0.004.

As shown in Spiegel (1963: 158), the estimated standard error for a proportion is se = ((p^*^q)/n)^5^. Applying this to the Gangelt data yields an estimated standard error of 0.00063, where 0.00063 = ((0.004^*^0.996)/9,957)^.5^. Multiplying this standard error by the z-score 1.96 and adding and subtracting this product to the estimated case fatality rate provides a 95% confidence interval for the latter, viz, 0.004 ± 1.96*0.00063. Thus, the estimated lower limit of the 95% confidence interval for the estimated case fatality rate of 0.004 is 0.00277 and the upper limit is 0.00524.

With this confidence interval in hand, we can translate it to the estimated number of unconfirmed, positive COVID-19 cases in the U.S. (4,639,790) found earlier by dividing the German case fatality rate (**R** =0.004) into the reported COVID-19 deaths (18,559). The translation follows work by Espenshade and Tayman (1982) on developing a confidence interval for a population estimate based on a censal-ratio method. However, we need to use a tolerance factor rather than a z score because we are not estimating the “mean” number of positive, unconfirmed case, but, rather, the total number. The appropriate tolerance factor for a 95% confidence interval is 2.036 (Espenshade and Tayman, 1982: 201-202), so for the lower limit of the case fatality rate we have 0.00272 and for the upper limit we have 0.00528. This leads to 3,513,179 and 6,823,162 as the estimated lower and upper 95% confidence limits, where 3,513,179 = (18,559 /0.00528) and 6,823,162 = (18,559/0.00272).

Recall the ratio of estimated unconfirmed cases to confirmed cases shown earlier, (*U*_i_/**C**_i_), where (*U*_i_/**C**_i_) = 9.4 = 4,639,790/492,416. We can now estimate its 95% confidence interval as being from 7.13 to 13.86, where (*U*_i_/**C**_i_)_ll95%_ = 7.13 = 3,513,179/492,416 and (*U*_i_/**C**_i_)_ul95%_ = 13.86 = 6,823,162/492,416.

Using the ratio of (*U*_i_/**C**_i_) = 9.4, we already found that the estimated number of positive, unconfirmed cases in Whatcom County as of April 25^th^ is 2,670, where *U*_w_ = 2,670 = 9.4^*^284. Using the procedure shown above to estimate the 95% confidence interval for the number of positive, unconfirmed cases in the U.S, we find that the 95% confidence interval for the estimate of *U*_w_ = 2,670 positive, unconfirmed cases in Whatcom County as of April 25^th^ is from 2,025 to 3,936, where U_wll95%_ = 2,025 = 7.13^*^284 and *U*_wul95%_ = 3,936 = 13.86^*^284.

We also can apply this to the number of positive, unconfirmed cases estimated by using the active cases instead of the total number of confirmed cases, viz, 2,416 = 9.4^*^ 257. The lower and upper 95% confidence intervals for the estimate that there are 2,416 positive, unconfirmed cases in Whatcom County as of April 25^th^ are 1,832 and 3,559, respectively, where 1,832 = 7.13^*^257 and 3,559 = 13.85^*^257.

Our approach to developing confidence intervals is an informal approximation to the “error propagation method” introduced by Deming (1950: 127-134). In different forms the error propagation method has been used by Alho and Spencer (2005), Espenshade and Tayman (1982), and Hansen, Hurwitz, and Madow (1953), among others. Swanson and Tayman (2014) found that the informal approach yields approximately the same results as the formal approach. So, we use it here to obtain a 95% confidence interval for the total number of positive, unconfirmed COVID-19 cases by, as we described earlier, adding the lower and upper boundaries of the intervals for the components leading up to the final estimate of positive, unconfirmed cases for Whatcom County.

### Concluding Remarks

It is clear that in using the case fatality rate from Gangelt, Germany (Bailey, 2020), we are making the assumption that it fits the U.S. and, ultimately, Whatcom County, Washington. We are forced to do this because no data comparable to the Gangelt study are yet available in the U.S. and where they are available, e.g., South Korea, (Shim et al., 2020), we believe that they are less suited to the U.S. than the Gangelt data because the containment measures, testing, and tracking, are less representative of the U.S. than the is the case with the Gangelt study. “Case fatality rates” in the U.S. are at least as suspect because they appear to be calculated in the absence of the total number of positive, unconfirmed cases.

## Data Availability

Data are available from public sources as cited in the article.

## Footnotes

* The authors are grateful to Jeff Tayman for comments.

